# Telehealth and Outpatient Utilization: Trends in Evaluation and Management Visits Among Medicare Fee-For-Service Beneficiaries, 2019-2024

**DOI:** 10.1101/2025.03.05.25323449

**Authors:** James D. Lee, Elena Chun, Chiang-Hua Chang, Terrence Liu, Rodney L. Dunn, Jeffrey S. McCullough, Michael P. Thompson, Chad Ellimoottil

**Affiliations:** Institute for Healthcare Policy and Innovation, University of Michigan; Department of Internal Medicine, University of Michigan; Department of Urology, University of Michigan; Department of Cardiac Surgery, University of Michigan; Department of Health Management and Policy, University of Michigan

**Author notes:** Corresponding Author: Chad Ellimoottil, MD, MS, Associate Professor, Department of Urology, University of Michigan, Institute for Healthcare Policy and Innovation, University of Michigan, 2800 Plymouth Road, Building 16, Ann Arbor, MI 48109.

## Abstract

**Introduction:** Telehealth expanded rapidly following the COVID-19 pandemic and has become an integral part of healthcare delivery. However, concerns remain that increased telehealth availability may contribute to higher overall healthcare utilization and spending. To assess telehealth’s impact on outpatient evaluation and management (E&M) visit volume, we compared overall E&M utilization before and after the pandemic across specialties with varying levels of telehealth use.

**Methods:** We analyzed 100% Medicare Fee-For-Service (FFS) claims to compare monthly outpatient E&M visit rates between two periods: pre-pandemic (January 2019-February 2020) and post-pandemic (January 2021-June 2024). Specialties were categorized by telehealth use as high (behavioral health), medium (primary care), and low (orthopedic surgery). A difference-in-differences (DID) analysis was used to assess changes in visit volume associated with telehealth.

**Results:** Prior to the pandemic, telehealth accounted for just 0.1% of monthly E&M visits but surged to 41.0% in April 2020 before stabilizing between 5.7% and 7.0% in 2023-2024. The average monthly E&M visit rate per 1,000 FFS beneficiaries was 906.8 pre-pandemic and 918.6 post-pandemic. In the post-pandemic period, telehealth comprised 1.2% of E&M visits in low-use specialties, 8.4% in medium-use specialties, and 43.8% in high-use specialties. Compared to the expected trend based on the low telehealth-use specialty, high and medium telehealth-use specialties experienced a 4.1% and 7.2% relative decline in overall E&M visits, respectively, in the post-pandemic period.

**Conclusion:** Following an initial surge, telehealth use stabilized in 2021 and beyond. Overall outpatient utilization remained stable post-pandemic, and increased telehealth adoption was not associated with a rise in total outpatient E&M visits. These findings suggest that broad telehealth adoption has not led to increased healthcare utilization among Medicare FFS beneficiaries.

## Introduction

Telehealth use expanded rapidly following the COVID-19 pandemic, serving as an essential alternative to in-person care.^1^ In response, the federal government implemented temporary Medicare telehealth coverage policies to support this expansion during the public health emergency.^1^ Although telehealth use declined after peaking in 2020, it has remained an integral part of healthcare delivery in the post-pandemic era, continuing to represent a substantial share of healthcare services.^2,3^

While there has been bipartisan support for telehealth, telehealth coverage extensions have been temporary.^4–6^ A primary barrier to permanent policy changes is the lack of clear evidence on telehealth’s impact on Medicare spending. In particular, concerns persist that expanded telehealth use may drive additional healthcare utilization, leading to increased Medicare spending.

As federal policymakers consider permanent telehealth coverage, it is essential to assess post-pandemic telehealth trends and whether expanded use has contributed to an increase in overall service volume. This study examines trends in outpatient evaluation and management (E&M) visits, which constitute a significant portion of Medicare spending.^7^ We first analyzed overall trends in E&M visit volume among Medicare beneficiaries before and after the pandemic. We then conducted an analysis comparing trends across three specialties with high, medium, and low telehealth use to assess whether telehealth use was associated with changes in outpatient visit volume.

## Methods

### Data and Population

We analyzed 100% of Medicare Fee-For-Service (FFS) Carrier and Outpatient claims from January 2019 to June 2024. Each year, we included only FFS beneficiaries with Part A and Part B coverage and no Medicare Advantage enrollment.

For the primary analysis, we excluded beneficiaries under 65 years of age to improve comparability across the three specialty groups and reduce potential bias from those eligible for Medicare due to disability or end-stage renal disease. Additionally, we excluded counties with fewer than 10 FFS beneficiaries or fewer than 10 E&M visits per month per specialty.

### Identifying Overall E&M Visits and Telehealth E&M Visits by Specialty

We identified all outpatient E&M visits during the study period using Berenson-Eggers Type of Service codes. Telehealth visits were classified based on Medicare’s list of eligible telehealth services and the corresponding modifier or place of service codes for each study year.^8^ Additionally, Healthcare Common Procedure Coding System (HCPCS) codes and revenue codes were used to identify telehealth services provided at Federally Qualified Health Centers and Rural Health Clinics.

Using Provider Specialty Codes, we categorized E&M visits into three groups:

- Low telehealth use (orthopedic surgery)
- Medium telehealth use (primary care)
- High telehealth use (behavioral health)

This classification was based on observed variations in telehealth adoption across specialties, with behavioral health providers historically utilizing telehealth at higher rates, primary care adopting it at moderate levels, and surgical specialties relying on it the least.^9^

### Outcomes

Our primary outcome was the monthly total E&M visits per 1,000 FFS beneficiaries. The numerator was the total number of E&M visits, including both in-person and telehealth, while the denominator was the number of eligible FFS beneficiaries enrolled in that month.

We then applied a model to compare changes in total E&M visits across specialties with high, medium, and low telehealth use. In this model, the count variable was the total number of E&M visits for each specialty as the numerator. To adjust for differences across counties, we included county-level fixed effects, which account for factors unique to each county that might influence healthcare use, such as local healthcare infrastructure or population demographics. Therefore, the denominator for the outcome in the model was the number of FFS beneficiaries in each county.

### Statistical Analysis

To assess how telehealth affected total E&M visits, we conducted a difference-in-differences (DID) analysis with county fixed effects. We defined two time periods: pre-pandemic (January 2019 – February 2020) and post-pandemic (January 2021 – June 2024). While the pandemic extended beyond January 2021, we use “post-pandemic” to refer to a period when telehealth use had stabilized. In contrast, March 2020 – December 2020 was a time of significant fluctuation in telehealth adoption, making it unsuitable for direct comparisons Rather than simply comparing total E&M visits before and after the pandemic, we examined trends across three specialties with low (orthopedic surgery), medium (primary care), and high (behavioral health) telehealth adoption. This approach creates a control group to account for factors unrelated to telehealth that could affect E&M visits.

For example, if Medicare policy changes had increased reimbursement for outpatient visits, we might see a rise in total E&M visits regardless of telehealth use. Similarly, if patients delayed care during the height of the pandemic and then sought more in-person visits later, this could also contribute to an increase in E&M visits, independent of telehealth availability.

To isolate telehealth’s impact, we used a low telehealth adoption specialty (orthopedic surgery) as a baseline to represent the general trend in outpatient visits. We then compared changes in total E&M visits before and after the pandemic for this low telehealth specialty against two specialties with medium and high telehealth adoption.

If telehealth were the primary driver of increased total E&M visits, we would expect specialties with higher telehealth use to experience a greater rise in total visit volume than the specialty with low telehealth use. Notably, our analysis confirmed that total E&M visit rates were parallel across all three specialties before the pandemic, a key assumption required for a valid DID analysis.

## Results

### Telehealth Use

The average monthly rate of telehealth E&M visits was 1.1 per 1,000 beneficiaries (0.1% of total E&M visits) in 2019 but surged during the pandemic, peaking at 231.3 per 1,000 beneficiaries (41.0%) in April 2020 (Figure 1). Telehealth use then gradually declined, averaging 86.0 (9.7%) in 2021, 67.5 (7.5%) in 2022, 57.9 (6.2%) in 2023, and 57.6 (6.0%) in 2024.

**Figure 1.**
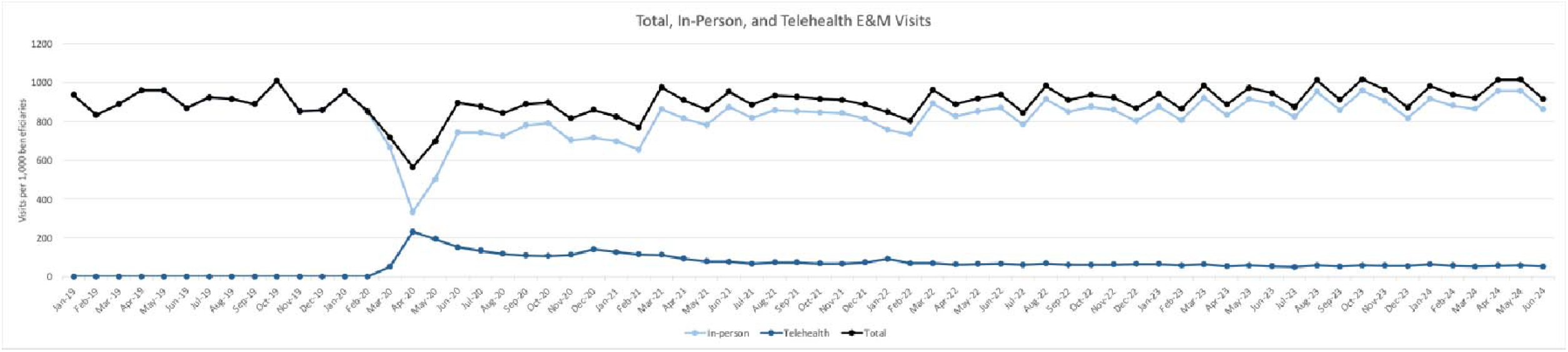
Monthly Trends in Outpatient Evaluation and Management (E&M) Visits per 1,000 Beneficiaries in Medicare Fee-for-Service, 2019–2024

For orthopedics (low telehealth use specialty), telehealth visits were rare before the pandemic, with 0.001 per 1,000 beneficiaries (0.004%) in 2019 (Figure 2). Usage peaked at 2.4 per 1,000 beneficiaries (20.4%) in April 2020, then declined to 0.4 (1.5%) in 2021 and remained stable at 0.3 (1.1%) in 2022, 1.0% in 2023, and 1.2% in 2024.

**Figure 2.**
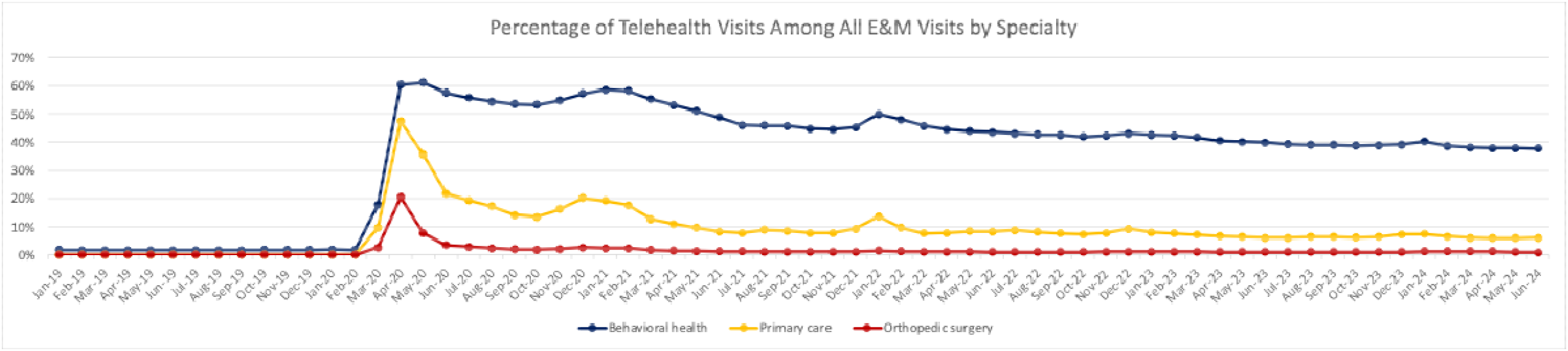
Monthly Trend in Percentage of Telehealth Visits Among All Evaluation and Management (E&M) Visits for Behavioral Health, Primary Care, and Orthopedic Surgery for Medicare Fee-for-Service Beneficiaries, 2019-2024.

For primary care (medium telehealth use specialty), telehealth visits were also low in 2019, averaging 0.04 per 1,000 beneficiaries (0.03%) (Figure 2). The rate peaked at 45.0 (47.3%) in April 2020, then gradually declined but remained higher than orthopedics, averaging 12.7 (10.7%) in 2021, 10.1 (8.7%) in 2022, 7.8 (6.7%) in 2023, and 7.4 (6.3%) in 2024.

For behavioral health (high telehealth use specialty), telehealth visits followed a similar trajectory, with 0.5 per 1,000 beneficiaries (1.6%) in 2019 (Figure 2). Usage increased sharply to 16.3 (57.2%) in December 2020, then declined but remained significantly higher than primary care and orthopedics, averaging 13.5 (49.7%) in 2021, 11.1 (49.7%) in 2022, 9.9 (40.0%) in 2023, and 9.3 (38.4%) in 2024.

### Total E&M Visits

Before the pandemic, the monthly rate of outpatient E&M visits per 1,000 FFS beneficiaries ranged from 832.1 (February 2019) to 1,008.1 (October 2019) (Figure 1). The rate dropped sharply at the onset of COVID-19, reaching a low of 564.5 in April 2020, before gradually recovering to 895.3 in 2021, 901.4 in 2022, 936.8 in 2023, and 963.4 in 2024.

For the low telehealth use specialty, the average monthly total E&M visit rate per 1,000 beneficiaries was 27.3 in 2019, declining to a low of 11.6 in April 2020 (Figure 3). The rate then rebounded to 25.9 in 2021 and remained stable at 26.1 in 2022, 26.9 in 2023, and 27.3 in 2024.

**Figure 3.**
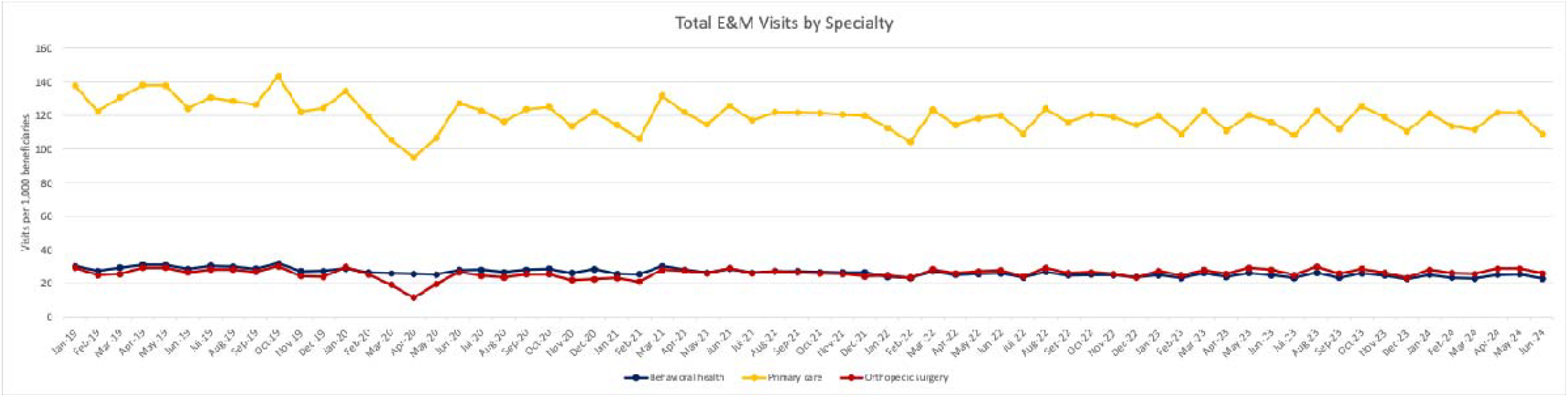
Monthly Trend in Outpatient Evaluation and Management (E&M) Visits per 1,000 Beneficiaries for Behavioral Health, Primary Care, and Orthopedic Surgery in Medicare Fee-for-Service, 2019-2024.

For the medium telehealth use specialty, the average monthly visit rate was 130.6 per 1,000 beneficiaries in 2019, dropping to 95.0 in April 2020 (Figure 3). The rate then increased to 119.8 in 2021 and remained relatively stable at 116.4 in 2022, 116.5 in 2023, and 116.5 in 2024.

For the high telehealth use specialty, the average monthly visit rate was 29.4 per 1,000 beneficiaries in 2019, followed by a gradual decline during and after the pandemic (Figure 3). The rate averaged 27.2 in 2021, 25.1 in 2022, 24.7 in 2023, and 24.2 in 2024.

### Impact of Telehealth on Outpatient Visit Volume: Model Results

#### Pre-Pandemic Period (January 2019 – February 2020)

Before the pandemic, behavioral health had 27% fewer total E&M visits per beneficiary at baseline compared to orthopedics (Table). In contrast, primary care had a significantly higher baseline visit rate than orthopedics, with 4.76 times more visits per beneficiary.

**Table 1.** Difference-in-Differences Model Estimate for the Association of Specialties with High, Medium, and Low Telehealth Use with Rates of Outpatient Evaluation and Management (E&M) Visits per 1,000 Beneficiaries at the County-Level.

| Variable | Coefficient | Rate Ratio (Exp(B)) | 95% CI | p-value |
| --- | --- | --- | --- | --- |
| Post-Pandemic Period | -0.0379 | 0.9628 | (0.9601, 0.9655) | <0.001 |
| Behavioral Health (BH) Specialty | -0.3181 | 0.7275 | (0.7212, 0.7339) | <0.001 |
| Primary Care (PC) Specialty | 1.5613 | 4.7651 | (4.7274, 4.8032) | <0.001 |
| BH x Post-Pandemic | -0.0415 | 0.9593 | (0.9543, 0.9644) | <0.001 |
| PC x Post-Pandemic | -0.0752 | 0.9276 | (0.9215, 0.9337) | <0.001 |

#### Pre-vs. Post-Pandemic Trends (January 2021 – June 2024)

The post-pandemic period was associated with a slight overall decline in total E&M visits (Table). The low telehealth-use group (orthopedics) experienced a 3.7% reduction in total visit volume compared to the pre-pandemic period. In the medium telehealth use group (primary care), E&M visit volume was 7.2% lower than projected, while in the high telehealth use group (behavioral health), it was 4.1% lower, both relative to trends in orthopedics.

## Discussion

This study examined the impact of telehealth adoption on outpatient E&M visit volume among Medicare FFS beneficiaries across three specialties with varying levels of telehealth use: orthopedic surgery (low), primary care (medium), and behavioral health (high). We found that telehealth was widely adopted in mental health and primary care compared to orthopedic surgery, with usage peaking at the onset of the pandemic before stabilizing from 2021 to 2024. Despite this sustained telehealth adoption, total outpatient E&M visits remained relatively unchanged or slightly declined, suggesting that telehealth primarily substituted for in-person visits rather than increasing overall healthcare utilization.

Several factors may explain why total E&M visit volume did not increase despite the expanded availability of telehealth. First, provider capacity remained a limiting factor—most clinicians operate on fixed schedules, and telehealth did not necessarily expand appointment availability. Second, the ongoing shortage of primary care and mental health providers may have constrained the ability to accommodate additional visits, even with telehealth. Third, some patients may have used telehealth selectively for convenience (e.g. reduced driving distance or time away from work) rather than increasing their overall demand for care.

These findings suggest that concerns about telehealth driving excessive healthcare utilization may be overstated, particularly in Medicare FFS populations. As policymakers consider permanent telehealth reimbursement policies, the evidence indicates that telehealth is largely substitutive rather than additive to in-person care, supporting its role as an alternative care delivery model rather than a driver of increased utilization.

This study has several limitations. First, we used Medicare FFS data, so the findings may not be generalizable to beneficiaries with Medicare Advantage or commercial insurance. However, understanding telehealth’s impact on Medicare FFS utilization and spending is crucial for informing permanent telehealth policy. Second, while the DID approach accounts for many unobserved differences between specialties, time-varying confounders may still bias our findings. Third, this study does not evaluate the quality of telehealth visits compared to in-person E&M visits. Fourth, while E&M visits represent a significant component of healthcare utilization, we did not account for other areas of healthcare use, such as diagnostic testing, imaging, or procedures.

## Conclusion

Compared to the low telehealth use specialty, high and medium telehealth use specialties experienced a slight decline in overall outpatient E&M visits. These findings suggest that the widespread adoption of telehealth post-pandemic did not lead to an increase in total outpatient E&M utilization among Medicare FFS beneficiaries. Instead, outpatient telehealth services appear to have primarily substituted for, rather than added to, in-person visits.

## Data Availability

All data in the study are publicly available through the Chronic Conditions Data Warehouse (www2.ccwdata.org).

